# Screening for adverse childhood experiences in pediatric clinical settings: a scoping review

**DOI:** 10.1101/2024.10.21.24315896

**Authors:** María José Correa-Méndez, Mariana Vásquez, Laura Restrepo, Valeria Diaz, Cristina Escobar, Isabella Franky, Isabela Arango, Oscar Gómez, Lina Maria Gonzáles Ballesteros

**Affiliations:** Medical student, Faculty of Medicine, Pontificia Universidad Javeriana, Bogotá, Colombia; Department of Psychiatry and Mental Health, Faculty of Medicine, Pontificia Universidad Javeriana, Bogotá, Colombia; Psychiatrist, PhD(c), Clinical Epidemiology, Professor in the Department of Psychiatry and Mental Health, Faculty of Medicine, Pontificia Universidad Javeriana. Leader of Physical and Socioemotional Well-being, Saldarriaga Concha Foundation, Bogotá, Colombia

**Keywords:** Perceptions, barriers, screening tools, adverse childhood experiences (ACE), screening

## Abstract

**Objective:** To compile the available literature regarding the usefulness, methodology used, and limitations of ACEs screening in a pediatric clinical sample, to prevent adverse health outcomes.

**Introduction:** Several studies have found an association between the presence of adverse childhood experiences (ACEs) and the development of negative health outcomes, however the usefulness of screening for this condition and how to do so appropriately has not been established.

**Methods:** We included observational, descriptive, analytical, and experimental studies, qualitative studies, systematic reviews, clinical guidelines, and health policies that include the pediatric population from 3 to 17 years of age in a clinical setting. Studies in languages other than Spanish and English, protocols, and validation studies were excluded.

The search strategy was designed using the PRISMA-S protocol to locate published and unpublished studies using PubMed, EMBASE, Cochrane, LiLACS, PsycArticles and CINAHL databases and grey literature in ProQuest Dissertations and Theses Global. Studies in Spanish and English published since 2012 were included. The Rayyan platform was implemented to remove duplicates and for independent article selection by each author, with conflicts resolved through discussion. Data was extracted into an Excel database, highlighting information related to the objectives of the scoping review. After analyzing the data, we present them in different tables that summarize the findings of the review, with a narrative description.

**Results:** Our search yielded 8143 studies, from which 31 studies met the inclusion criteria. These studies varied in design, with most conducted in the United States, they were published between 2012 and 2024. The review identified several ACE screening tools (15), with an average response time of 15 minutes. Although some tools showed promise in identifying children at risk for adverse outcomes, significant gaps remain in the consistency and effectiveness of screening methods. Barriers such as lack of training for healthcare personnel, a lack of approach to patients with positive results, limited resources for follow-up care, and cultural differences in interpreting ACEs were often highlighted. There was no evidence of adequate standardization of screening methods, how to use them, and how to properly identify and categorize the presence of an ACE in childhood.

**Conclusions:** This scoping review highlights the potential of screening for ACEs in pediatric settings to prevent long-term health issues. However, the lack of standardized tools, protocols, and evidence of long-term benefits hinders its implementation. Although the association between ACEs and negative health outcomes is well-established, there is insufficient evidence demonstrating the effectiveness of screening. Cultural factors, especially in countries like Colombia, further complicate the adaptation of screening tools. The review suggests that a multidisciplinary, family-centered approach and training in trauma-informed care are essential. More research is needed to standardize ACE screening and evaluate its effectiveness in reducing health issues related to ACEs.

## Introduction

An adverse childhood experience (ACE) is a potentially traumatic event or psychosocial factor experienced by a child before turning 18 (1). Since being coined in 1998, they have been classified under 3 main domains: abuse, neglect, and household dysfunction, defined together as infliction of physical, sexual, or emotional harm through the acts of an individual’s caregiver (2). Each of these domains has subcategories, gathering a total of 10 categories of ACEs (1). ACEs are highly prevalent, affecting 52% of children, with parental substance abuse being the most frequently reported experience. Females tend to report more ACEs than males, and non-white race/ethnicity, low education and low socioeconomic status have been significantly associated with reporting ACEs (3,4).

ACEs have been previously linked to a wide array of adverse health outcomes, including risky health behaviors, chronic health conditions, low quality of life, mental illnesses, and early death (3). Its negative impact on health is not only evident during childhood, but in adulthood as well. In children and adolescents, they have been associated with health outcomes such as asthma, sleep disturbances, recurrent infections, and cognitive delays, whereas on the social sphere they have been linked to delinquent behavior, bullying, dating violence and weapon carrying (3,5). On the other hand, there has been strong dose-response relationship between the number of ACEs and type 2 diabetes, cardiovascular disease, chronic lung disease, autoimmune disorders and sleep disturbances, and other outcomes regarding mental health such as depression, anxiety, substance abuse, post-traumatic stress disorder and violence in adults (1,3).

According to the above, adequately screening for ACEs in theory would allow for identification of at risk individuals for early interventions, which could mean a decrease in health-related outcomes (6); however, there are studies in the literature that analyze the possibility that screening for ACEs doesn’t have a significant effect on health outcomes (7). Therefore, we consider that it is important to assess which are the current screening strategies available.

### Objective and review question

This scoping review was conducted to summarize the available literature regarding the tools, usefulness, recommendations, perceptions and gap of knowledge of screening for ACEs in a pediatric clinical setting to prevent adverse health outcomes. The question we formulated was: is screening for adverse childhood experiences (ACEs) in the pediatric population in a clinical setting considered useful to prevent undesired health outcomes, how can it be done effectively, what are the limitations surrounding it and what are the tools available for doing so?

### Inclusion criteria

#### Participants

Children between 3-17 years of age.

#### Concept

Adverse childhood experiences screening, how to do it, tools for doing so, its importance, gaps in knowledge, and barriers.

#### Context

Clinical setting

#### Types of sources

This scoping review includes experimental studies, analytical observational studies such as prospective and retrospective cohort studies, and analytical cross-sectional studies, as well as descriptive observational and descriptive cross-sectional studies. Qualitative studies focusing on methodologies such as phenomenology, grounded theory, and qualitative description were also considered. Additionally, systematic reviews, literature reviews, clinical guidelines, user guides, and political contexts related to healthcare policies in different countries that meet the inclusion criteria were included. However, clinical protocols and scoping reviews were excluded.

### Exclusion criteria

- Studies not available in English or Spanish
- Full text not available
- Tools validation studies
- Scoping reviews or clinical protocols
- Community-based screening related articles

## Methods

Protocol and registration: This review was conducted in accordance with an *a priori* protocol with JBI Evidence Synthesis template (8). The final protocol was registered prospectively with the Open Science Framework in 2024.

### Search strategy

The search strategy is presented based on the PRISMA-S extension to the PRISMA Statement for Reporting Literature Searches in Systematic Reviews (9) It aims to locate both published and unpublished studies. An initial limited search of PubMed was undertaken to identify relevant articles on the topic. A search strategy with an objective approach was constructed by a health professional with experience on information retrieval (OG). MeSH, EMTREE and free terms for the major topics important to the review were selected from the controlled language thesaurus of the PubMed, EMBASE and PsycNet databases. Additional terms were identified using the online tools PubMed PubReMiner [MC1] (10) and MeSH on Demand (11), and from the abstracts of relevant papers previously known to the authors. See **Appendix I** for the MEDLINE (PubMed) and EMBASE (Elsevier) search strategy.

The following databases were be searched: MEDLINE (PubMed), EMBASE (EMBASE), Cochrane Central Register of Controlled Trials (Ovid), LiLACS (BVS), PsycArticles (PsycNet), CINAHL (EBSCO). The reference list of all included sources of evidence was screened for additional studies. Studies published in Spanish and English were included as the investigators are fluent in both languages. Studies published since 2012 were included as the topic is new and new information on ACEs screening is recent. The gray literature search was conducted in ProQuest Dissertations and Theses Global (ProQuest).

### Study/Source of evidence selection

Following the search, all identified citations were uploaded into Rayyan (12), and duplicates removed using the duplicate detection tool from the system and manual verification. Following a pilot test, titles and abstracts were screened by two or more independent reviewers for assessment against the inclusion criteria for the review. The full text of selected citations was assessed in detail against the inclusion criteria by two or more independent reviewers. Reasons for exclusion of sources of evidence at full text that do not meet the inclusion criteria were recorded and reported in the scoping review. Any disagreements that arose between the reviewers at each stage of the selection process was resolved through discussion, or with an additional reviewer/s. The results of the search and the study inclusion process was reported in full in the final scoping review and presented in a Preferred Reporting Items for Systematic Reviews and Metaanalyses extension for scoping review (PRISMA-ScR) flow diagram (13).

### Data extraction

Data was extracted from papers included in the scoping review by all independent reviewers using a data extraction sheet developed by the reviewers in Microsoft Excel. The data extracted included name of the article, authors, location, year in which it was published, dates in which the study was made, population specifics, type of study, objective of study, used tools, strategies to screen correctly, outcomes that could be prevented, perceptions surrounding screening, barriers and gaps of knowledge. The draft data extraction tool was modified and revised as necessary during the process of extracting data from each included evidence source. Any disagreements that arose between the reviewers were resolved through discussion, or with an additional reviewer/s.

### Data analysis and presentation

The results are presented in several formats. First, a PRISMA flow diagram (**Figure 1)** illustrates the selection process, accompanied by a paragraph summarizing the included articles. **Table 1** describes the key characteristics of the included studies, including the author(s), title, location, year of publication, and study design. **Table 2** provides a summary of the screening tools identified, including details such as the country of origin, the country where the tool was validated, administration method, target population, number of items, content of the items, time required to complete, response options, scoring system, and information on reliability and validity. A narrative summary follows, outlining the undesired health outcomes that may be prevented by using the screening tools and the overall advantages of screening. Another narrative summary details the recommended strategies for effective implementation of the screening tools in clinical practice, as identified through the review. Lastly, a concluding section summarizes the perceptions of parents and medical staff regarding screening, the existing knowledge gaps, and the perceived barriers to implementing screening practices.

**Table 1:** Description of included studies. **Supplemental file

| Author | Title | Location | Year of publication | Study design |
| --- | --- | --- | --- | --- |
| Jacob et al. | Adverse childhood experiences: Basics for the Paediatrician | Canada | 2018 | Narrative review |
| Sherfinski et al. | Adverse Childhood Experiences: Perceptions, Practices, and Possibilities | United States of America | 2021 | Narrative review |
| Marie-Mitchell et al. | Adverse childhood experiences: translating knowledge into identification of children at risk for poor outcomes | United States of America | 2012 | Cross-sectional study |
| Garton et al. | A national survey on the use of screening tools to detect physical child abuse | Germany | 2016 | Cross-sectional study |
| Reading et al. | A Qualitative Study of Pediatricians' Adverse Childhood Experiences Screening Workflows. | United States of America | 2022 | Qualitative study based on semi-structured interviews |
| Negriff et al. | Assessment of Screening for Adverse Childhood Experiences and Receipt of Behavioral Health Services Among Children and Adolescents. | United States of America | 2022 | Retrospective cohort study |
| Vega-Arcea et al. | Cribado de las experiencias adversas en la infancia en preescolares: revisión sistemática | Chile | 2017 | Systematic review |
| Koital et al. | Development and implementation of a pediatric adverse childhood experiences (ACEs) and other determinants of health questionnaire in the pediatric medical home: A pilot study. (BARC Pediatric Adversity and Trauma Questionnaire) | United States of America | 2018 | Pilot study |
| Barnes et al. | Identifying adverse childhood experiences in pediatrics to prevent chronic health conditions. | United States of America | 2020 | Narrative review |
| Watson et al. | Implementation and Evaluation of Adverse Childhood Experiences Screening in Pediatrics and Obstetrics Settings. | United States of America | 2024 | Pilot study |
| Hillyer | Implementation of Pediatric Adverse Childhood Experiences Screening in Primary Care | United States of America | 2021 | Observational descriptive pilot study |
| Marie-Mitchell et al. | Implementation of the Whole Child Assessment to Screen for Adverse Childhood Experiences | United States of America | 2019 | Observational descriptive pilot study |
| Bora et al. | Inter-Clinician Variability in Primary Care Providers' Adverse Childhood Experience Knowledge, Training, Screening Practices, and Perceived Intervention Barriers: an Exploratory Cross-Sectional Study | United States of America | 2021 | Cross-sectional study |
| Bethell et al. | Methods to Assess Adverse Childhood Experiences of Children and Families: Toward Approaches to Promote Child Well-being in Policy and Practice | United States of America | 2017 | Systematic review |
| Rahman et al. | On Becoming Trauma-Informed: Role of the Adverse Childhood Experiences Survey in Tertiary Child and Adolescent Mental Health Services and the Association with Standard Measures of Impairment and Severity | Canada | 2018 | Cross-sectional study |
| Conn et al. | Parental perspectives of screening for adverse childhood experiences in pediatric primary care | United States of America | 2017 | Qualitative design |
| Liu et al. | Pediatric ACES assessment within a collaborative practice model: Implications for health equity | United States of America | 2021 | Mixed-methods |
| Bright et al. | Primary Care Pediatricians' Perceived Prevalence and Surveillance of Adverse Childhood Experiences in Low-Income Children. | United States of America | 2015 | Cross-sectional study |
| Finkelhor | Screening for adverse childhood experiences (ACEs): Cautions and suggestions. | United States of America | 2017 | Critical review |
| McBain et al. | The effect of adverse childhood experience training, screening, and response in primary care: a systematic review | United States of America | 2023 | Systematic review |
| DiGangi & Negriff | The Implementation of Screening for Adverse Childhood Experiences in Pediatric Primary Care. | United States of America | 2020 | Retrospective descriptive observational study |
| Cibralic et al. | Utility of screening for adverse childhood experiences (ACE) in children and young people attending clinical and healthcare settings: a systematic review. | Australia | 2022 | Systematic review |
| Bucci et al. | ACE-Q: User Guide for Health Professionals | United States of America | 2015 | User-guide |
| Dube | Continuing conversations about adverse childhood experiences (ACEs) screening: A public health perspective | United States of America | 2018 | Narrative review |
| Kia-Keating et al. | Trauma-Responsive Care in a Pediatric Setting: Feasibility and Acceptability of Adverse Childhood Experiences (ACEs) Screening | United States of America | 2020 | Interventional study |
| Kerker et al. | Do pediatricians ask about adverse childhood experiences in pediatric primary care? | United States of America | 2016 | Cross-sectional study |
| Albaek et al. | Walking children through a minefield: how professionals experience exploring adverse childhood experiences. | United States of America & Netherlands | 2018 | Qualitative metasynthesis |
| Bethell et al. | Methods to assess adverse childhood experiences of children and families: toward approaches to promote child well-being in policy and practice. | United States of America | 2017 | Cross-sectional study |
| Oh et al. | Review of tools for measuring exposure to adversity in children and adolescents | United States of America | 2018 | Systematic review |
| Garner et al. | Early childhood adversity, toxic stress, and the role of the pediatrician: translating developmental science into lifelong health. | United States of America | 2012 | Policy statement |
| Campbell | Screening for adverse childhood experiences (ACEs) in primary care: a cautionary note | United States of America | 2020 | Critical review |

**Table 3.** Summary of findings by key questions

| Outcome | Findings |
| --- | --- |
| Q1: Undesired health outcomes that might be prevented using screening tools and the overall advantages of screening | - ACEs are linked to PTSD, anxiety, depression, behavioral issues, cardiovascular disease, chronic lung disease, cancer, autoimmune diseases, diabetes, obesity, and sleep disturbances.<br>- Toxic stress from ACEs leads to health, behavioral, and social challenges, and a shorter life expectancy.<br>- Screening in primary care can identify families at risk, enable early intervention, and provide resources to mitigate these risks.<br>- Early screening can disrupt the progression of ACEs outcomes and break the intergenerational cycle of trauma, improving overall health and behavior. |
| Q2: Recommended strategies for the adequate employment of the tools in the clinical setting found during the search | - Choose screening tools based on patient age, condition, and setting.<br>- Lack of provider training is a major obstacle, reducing confidence and effectiveness in ACEs screening.<br>- Training healthcare professionals in validated tools is essential for meaningful outcomes.<br>- Implement a multidisciplinary, multicultural, and family-centered approach; provide clear context to parents and children for better accuracy.<br>- Familiarize the entire team with the screening system to ensure efficiency.<br>- Pediatricians play a key role in advising on nutrition, sleep, and other toxic stress factors.<br>- Address stigmatization to manage positive screens effectively and improve cooperation and understanding. |
| Q3: Perceptions of parents and medical staff regarding screening, the existing knowledge gaps, and the perceived barriers to implementing screening practices | - Healthcare providers and parents show varied perceptions of ACEs screening.<br>- Common barriers include a lack of awareness, insufficient knowledge, and unclear protocols.<br>- Many providers lack formal training and feel unprepared to discuss traumatic experiences, fearing harm.<br>- Ethical concerns arise when risks are identified but follow-up interventions are lacking.<br>- Limited resources and time constraints are significant challenges.<br>- Literacy, language, and cultural considerations are crucial to developing effective screening tools.<br>- Despite ACEs' association with negative outcomes, there is limited evidence that screening improves health outcomes on its own. |

**Figure 1:**
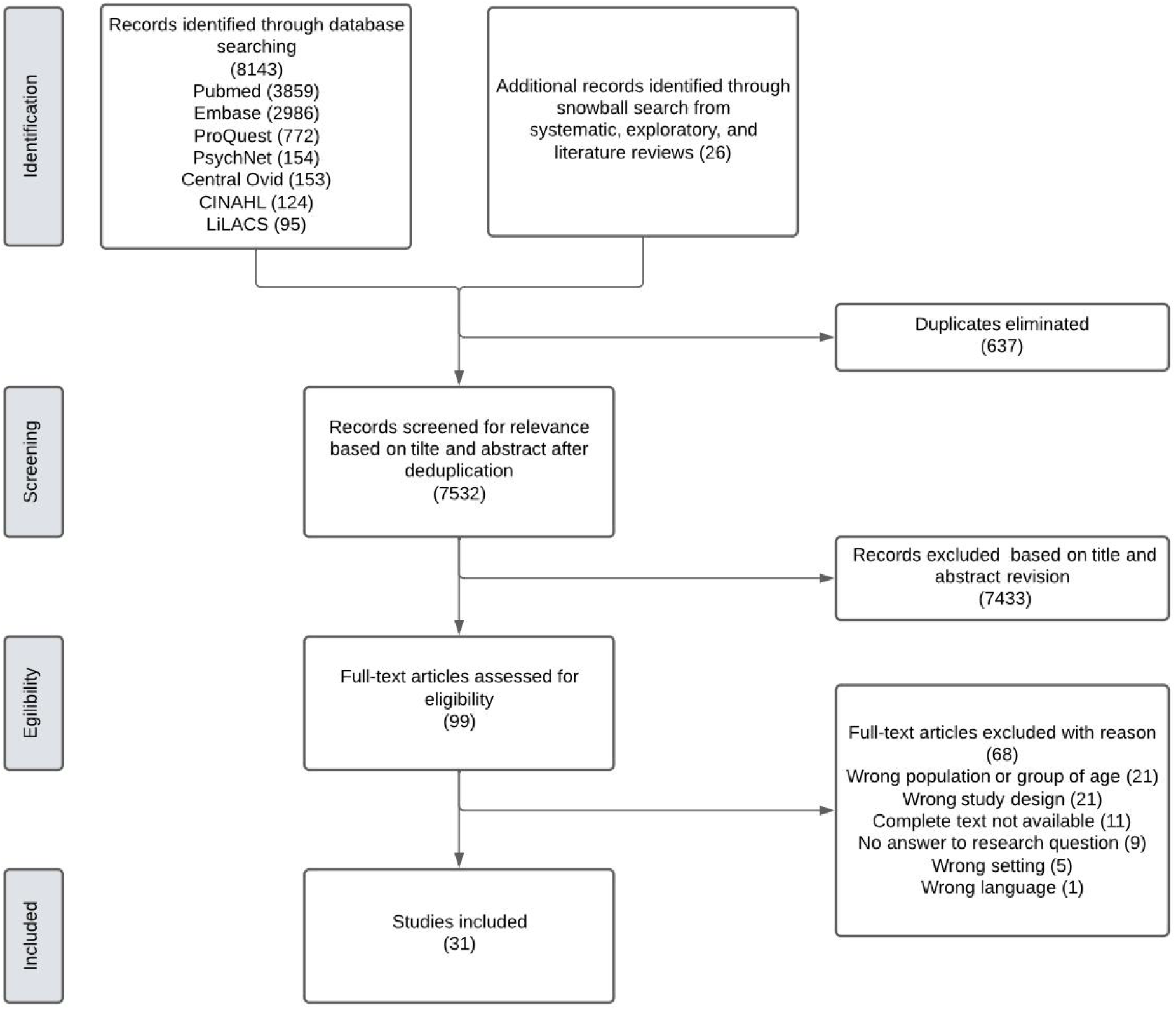
PRISMA flow diagram of study selection.

## Results

The search was carried out from February to September 2024 across seven databases, which yielded 8143 studies. Moreover, 26 additional records were identified with snowball search from systematic, exploratory, and literature reviews. We removed 637 duplicates and reviewed the 7532 titles and abstracts remaining, from which we excluded 7433. We retained 99 studies for full-text review, and 68 additional studies were excluded as they did not satisfy the inclusion criteria. Finally, 31 studies were included regardless of study quality to assure an exhaustive review (Figure 1).

These papers were published between 2012 and 2024, 14 being published in the past 5 years. Most studies were conducted in the United States of America (n=26), one of which was carried out in joint-work with the Netherlands. Only 2 were conducted in Canada, and 1 in Chile, Germany, and Australia respectively. 18 studies applied a quantitative design, 3 used qualitative methods, and 2 mixed methods. 8 were other kinds of manuscripts, which included 4 narrative reviews, 2 critical reviews, 1 user guide and 1 policy statement (Table 1).

**Table 2: Summary of the screening tools identified**

This table summarizes 15 screening tools, most of them were developed and validated in the United States, but seven of these tools have been validated in other countries, including the United Kingdom, Spain, Portugal, Brazil, Germany, South Africa, and China among others. The administration methods for the screening tools vary with some involving interviews with parents, caregivers, and children, while others are in the form of questionnaires, reports and self-reports. The age range spans from 0 to 19 years, depending on the type of the tool. Two of the tools include different versions depending on the age group and some other tools incorporate both the parents and the child’s perspective. The topics assessing these tools include exposure to violence, discrimination, physical abuse, emotional abuse, domestic violence, access to healthcare, among others. On average each tool covers 15 topics with the number of topics ranging from a minimum of 7 to a maximum of 35 items. The average time required to complete the screening is on average 15 minutes, ranging from 5 minutes to 60 minutes of completion time. The majority of these tools use yes/no questions, some include frequency based responses and others allow for concise and/or more extended open responses.

### Undesired health outcomes that might be prevented using screening tools and the overall advantages of screening

As it was previously mentioned, exposure to ACEs has been associated with multiple detrimental health repercussions, impacting on physical and emotional health, thus including post-traumatic stress disorder, anxiety, depression and a wide array of behavioral issues. It was also found a strong dose-response association with cardiovascular disease, chronic lung disease, cancer, autoimmune disease, diabetes, obesity and sleep disturbances (14), while also being involved in the development of toxic stress, culminating in an overall cluster of health, behavioral and social challenges (15), and a shortened life expectancy.

In light of the previous information, these detrimental outcomes might be prevented by implementing ACEs screening on primary care facilities, as it can help identify families at risk for the negative health and developmental outcomes associated with toxic stress, allowing for early intervention and access to resources that can mitigate these risks (16). In this way, the progression of ACEs outcomes can be interrupted, while also potentially resolving behavioral alterations from exposure to ACEs by breaking the intergenerational cycle that is caused by ACEs perpetuation (17), resulting in general improvement of health.

### Recommended strategies for the adequate employment of the tools in the clinical setting found during the search

Knowing the great impact of ACEs screening for constructing better health outcomes, it is crucial to establish multiple strategies for primary care providers to adequately employ the screening tools in a clinical setting. The first thing to consider is to know the wide range of screening tools available, where the choice of which tool to implement will depend mostly on the specific condition being screened for, the age of the patient and the setting in which the screening takes place (18). It is also crucial to acknowledge the primary obstacle of screening for ACEs, which is the lack of training for primary care providers and professionals, posing obstacles in the understanding of trauma-informed care practices and, by that, causing low confidence among professionals in conducting ACEs screening (19). Therefore, the main strategy could be training professionals to implement validated tools, which will be crucial to make a meaningful impact on the outcomes prevented by the ACEs screening (20).

It is also important to mention the great importance of a multidisciplinary, multicultural, multilingual team and a family-centered approach when screening for ACEs (21). Context and explanation of the questionnaire should be given to both parents and children who can understand, as this is reflected in better and more exact responses, resulting in an accurate collection of the information available (22). It is worth noting that familiarizing the whole team with the screening system is of utter importance, as this will help make the process increasingly agile. Once ACEs screening is instaurated, it is important to not overemphasize the ACEs score, and rather to consider the patient’s symptoms before carrying out any intervention (17). It is also worth noting the fundamental role of pediatricians in mitigating the impact of ACEs is done by advising families on nutrition, exercise, sleep habits, key components of toxic stress, and therefore, ACEs (23). While taking this into consideration, it is key not only for the health care providers but also for the patients to understand the focus of ACEs screening by addressing stigmatization (17). This works as a crucial way to cope with a positive screen, making it the first step in the course of action to go towards an improvement regarding ACEs health outcomes.

### Perceptions of parents and medical staff regarding screening, the existing knowledge gaps, and the perceived barriers to implementing screening practices

Medical staff and parents expressed diverse perceptions and encountered several barriers regarding the screening of ACEs. A consistent theme across multiple studies was the lack of awareness and insufficient knowledge among healthcare providers and guardians about the process and benefits of screening. Healthcare providers indicated the complexity and variability in screening protocols as barriers for implementation, particularly in clinical settings where standardized guidelines were unclear (24,25). Many healthcare providers report a lack of formal training on ACEs, and familiarity with ACEs screening practices remains low. Screening decisions are often influenced by beliefs about the relevance of addressing parenting behaviors and social-emotional development in pediatric care (26). Additionally, limited time and existing workload were frequently identified as significant difficulties by medical staff to screen for ACEs (27). Lack of proper training left many providers feeling unprepared or hesitant to engage in discussions about potentially traumatic experiences with their patients, fearing that they might cause further harm or distress (28). Healthcare providers also raised concerns about the ethical implications of screening without adequate follow-up interventions. In many cases, screening for ACEs revealed significant risk factors, yet the available resources to address these issues were either insufficient or non-existent (29). Other barriers identified include the need to incorporate literacy, language, and cultural considerations when developing and applying tools for ACEs screening. Variations in these factors can significantly affect the comprehension of screening items, potentially compromising response accuracy (22). ACEs include various traumatic events, each influencing health outcomes differently and requiring tailored interventions. While ACEs are linked to negative health outcomes, evidence is lacking that screening prevents these effects or improves outcomes (30)

## Discussion

Our results are largely consistent with those of previous studies, which indicate significant gaps in the standardization of screening for ACEs. One major theme that emerged is the lack of a clear, universally accepted definition of what constitutes an ACE. Many of the tools included in our review used terms like “adversity” or “trauma” interchangeably, leading to variations in how these experiences are measured. This lack of clarity not only affects the screening process but also contributes to inconsistencies in identifying and addressing ACEs. Moreover, the screening tools we reviewed often lacked rigorous testing and validation. Many had only been used in a single study, and several scales were designed to assess broader traumas rather than ACEs specifically. This highlights a significant gap in the availability of reliable and validated tools for ACE screening. Furthermore, it is unclear how to tailor screening based on the child’s age or whether certain tools are designed for use with parents or the patient directly. There is also no clear guidance on the appropriate age to apply screening tools directly to children.

Just like our hypothesis before beginning the project, we identified certain gaps in the use of ACE screening in clinical settings. A critical finding is the lack of information on the value and process of ACE screening. Health professionals often lack sufficient guidance on whether it is truly beneficial to screen for ACEs, and if so, the best methods for doing so. While there are many proposed screening strategies, none have been universally adopted, and institutional protocols are lacking. This absence of clear guidelines leaves healthcare workers uncertain about how to proceed when a child screens positive for ACEs. There is also the risk of overwhelming the healthcare system if every child who screens positive for ACEs is referred for trauma-informed care. Given the current limitations in both mental health resources and trauma care, such saturation could hinder the delivery of timely and effective interventions. After the realization of the project, we remain uncertain of the actual utility of ACE screening at this moment and lean towards skepticism about its current practical use.

One of the most concerning findings is the lack of robust evidence that ACE screening improves long-term health outcomes. While a dose-response relationship between the number of ACEs and negative health effects is well established, prospective studies that demonstrate the efficacy of ACE screening in preventing these outcomes are sorely lacking. The current approach appears more preventative than promotive, often failing to influence immediate medical management. This raises questions about the cost-benefit balance of routine ACE screening. Cultural considerations must also be factored into the screening process. Populations from different cultural backgrounds may interpret key life events such as marriage, divorce, and child-rearing differently. This is further complicated by language barriers, as the way screening questions interpretation can vary significantly based on the language and cultural context of the family. Stigma is another significant concern. A positive ACE screen may create an atmosphere of constant concern from healthcare providers and can inadvertently label children with a “heavy mark” that affects their future interactions with the healthcare system and even their own behavior. This could undermine the benefits of early intervention. It is crucial to recognize that the rights and well-being of children are influenced not only by the healthcare system but also by various social entities, the government, their families, and other key actors so a multidisciplinary approach is of essence.

Additionally, most of the scales and studies included in our review were developed in the United States and are therefore heavily influenced by American cultural norms. Applying these tools to different populations, such as those in Colombia, could lead to inaccurate assessments. The need for culturally sensitive tools is clear. In Colombia, there are no existing studies focused on ACEs screening, highlighting a significant research gap. Implementing ACEs screening should ideally occur in countries with sufficient economic resources dedicated to prevention, which poses a challenge for our country. While we have systems in place, such as those managed by the *Instituto Colombiano de Bienestar Familiar* (ICBF), these focus primarily on the most vulnerable populations, often leaving out large portions of the population who might also benefit from such interventions. Most ACE-related studies are conducted in large urban centers within first-world countries, and there is a notable absence of research in rural areas and lower-income nations like Colombia. Specifically, in rural, peripheral areas of Colombia, where socioeconomic conditions are poor, implementing ACEs screening would be particularly difficult due to the concentration of health infrastructure—hospitals, doctors, and other resources—mainly in urban centers.

The majority of the studies we included were literature reviews, systematic reviews, and cross-sectional studies. It is important to note that this subject requires a more subjective approach, as there are no established protocols or a robust body of evidence to guide ACEs screening in clinical practice. Many studies we initially reviewed did not align with our inclusion criteria, which made the selection process challenging and limited the number of articles we could include. However, our study addressed an uncommon but crucial topic: the correct strategies for ACEs screening. We identified significant gaps in knowledge, particularly regarding how to effectively conduct the screening process and manage a positive screen. It would be highly valuable to conduct future studies focusing on the specific steps to follow after a positive screen and the best management practices for these patients. Numerous gaps remain, such as how to standardize ACEs screening across different contexts, determining the most effective tool, assessing whether widespread screening is beneficial, and understanding the legal implications of implementing such a system. Additionally, any potential screening system would require adjustments in the political and social frameworks of the country. More high-quality, evidence-based studies are needed to answer these questions and guide future practice. Moving forward, we would benefit from more diverse research methodologies, including clinical trials, retrospective studies, probability and clinical correlation studies, and prospective studies, to provide stronger, evidence-based guidance for ACEs screening.

### Strengths and Limitations

One of the key strengths of our study lies in the comprehensive search across multiple databases, ensuring that we gathered as much relevant information as possible. In addition, we included a wide variety of study types to thoroughly assess the available literature. By not excluding studies based on their quality, we aimed to provide a realistic overview of the current research landscape. Our rigorous review process involved screening 8,143 articles and narrowing them down to 31, which demonstrates the care and precision taken throughout. However, certain limitations were evident. Many studies lacked sufficient methodological details, such as exact dates, locations, or population characteristics. Additionally, some studies had small sample sizes or did not include control groups, limiting their overall robustness. Only a small subset of articles fully met our inclusion criteria, making the selection process challenging. In retrospect, more specific exclusion criteria could have streamlined the selection and review process.

### Biases

Observer bias was present as the selection of articles was influenced by the subjective understanding of what constitutes an ACE, although this was mitigated by the involvement of a large evaluation team that helped control for bias. Incomplete information bias may have arisen due to the inclusion of studies that did not provide comprehensive details on certain aspects, such as methodology or results, limiting the reviewers’ ability to assess the quality or relevance of the studies accurately. Additionally, publication bias may have influenced our review, as studies with positive or statistically significant results tend to be published more frequently, while those with negative or null results are less likely to be included.

## Conclusions

We consider that this scoping review highlights the significant potential of screening for ACEs in pediatric clinical settings to potentially prevent long-term health outcomes. However, current limitations in standardized tools, clear protocols, and evidence supporting long-term benefits hinder the widespread implementation of ACE screening. Although the association between ACEs and negative health outcomes is well-established, there is insufficient evidence demonstrating the effectiveness of screening in preventing these outcomes. Additionally, cultural and contextual factors, particularly in countries like Colombia, present further challenges in adapting screening tools developed in high-resource settings.

The findings from this review suggest that effective ACE screening requires a multidisciplinary, family-centered approach, with a focus on training healthcare professionals in trauma-informed care practices. Addressing these gaps will require further research, including large-scale, longitudinal studies to evaluate the real-world impact of ACE screening, as well as the development of culturally sensitive tools tailored to diverse populations. Ultimately, while ACE screening holds promise as a preventive measure, further efforts are needed to standardize its use, ensure adequate follow-up interventions, and assess its true efficacy in reducing the burden of ACE-related health conditions.

## Data Availability

All data produced in the present study are available upon reasonable request to the authors

## Funding

This paper does not have external funding, it was carried out using the researchers’ own resources with the exception of access to databases, which is covered by the subscriptions of the investigators’ institution.

## Author contributions

The authors confirm contribution to the paper as follows: Study conception: MJC. Encouragement to investigate and supervision of this work: LMG. OG. Study design: OG. Developed the theory: LMG.

MJC. OG. All authors contributed to the writing of this manuscript. All authors reviewed and approved the final version of the manuscript.

## Conflicts of interest

The authors do not have any conflicts of interest to report.

## Ethics and dissemination

Since this study does not involve human participants, collection of primary data, or use of unpublished secondary data, formal ethical approval from a human ethics committee is not necessary. Findings were disseminated through professional healthcare networks, meetings with pertinent collaborators, conference presentations and a publication in a scientific journal.

## Appendices

## Appendix I: Search Strategies in MEDLINE and EMBASE

| Database | Search Words | Results |
| --- | --- | --- |
| MEDLINE Search Strategy | (child*[Title/Abstract] OR kid[Title/Abstract] OR kids[Title/Abstract] OR teen*[Title/Abstract] OR adolescent*[Title/Abstract] OR adolescence[Title/Abstract] OR juvenile*[Title/Abstract] OR "Child"[Mesh] OR "Adolescent"[Mesh]) AND (((("Adverse Childhood Experiences"[Mesh]) OR ("adverse childhood experience"[Title/Abstract] OR "adverse childhood experiences"[Title/Abstract]))) AND (("Mass Screening"[Mesh]) OR (tool*[Title/Abstract] OR instrument*[Title/Abstract] OR questionnaire*[Title/Abstract] OR inventor*[Title/Abstract] OR screen*[Title/Abstract] OR survey*[Title/Abstract] OR measure*[Title/Abstract]))) | 3,837 |
| EMBASE Search Strategy | ((('childhood adversity'/exp) OR ('adverse childhood experience',ti OR 'adverse childhood experiences',ti)) AND (('screening'/exp) OR (tool*,ti OR instrument*,ti OR questionnaire*,ti OR (inventor*,ti OR screen*,ti OR survey*,ti OR measure*,ti)) AND (('child'/exp) OR ('adolescent'/exp) OR (child*,ti OR kid,ti OR kids,ti OR teen*,ti OR adolescent*,ti OR adolescence,ti OR juvenile*,ti))) AND ([embase]/lim NOT ([embase]/lim AND [medline]/lim)) | 5,757 |

## Notes

### Competing Interest Statement

The authors have declared no competing interest.

### Clinical Protocols

https://osf.io/fs5pe/

## References

1. Gilgoff R, Singh L, Koita K, Gentile B, Marques SS. Adverse Childhood Experiences, Outcomes, and Interventions. Pediatr Clin North Am. 2020 Apr;67(2):259–73.

2. Adigun OO, Mikhail AG, Krawiec C, Hatcher JD. Abuse and Neglect. In: StatPearls [Internet]. Treasure Island (FL): StatPearls Publishing; 2024 [cited 2024 Oct 14]. Available from: http://www.ncbi.nlm.nih.gov/books/NBK436015/

3. Petruccelli K, Davis J, Berman T. Adverse childhood experiences and associated health outcomes: A systematic review and meta-analysis. Child Abuse Negl. 2019 Nov;97:104127.

4. Felitti VJ, Anda RF, Nordenberg D, Williamson DF, Spitz AM, Edwards V, et al. Relationship of childhood abuse and household dysfunction to many of the leading causes of death in adults. The Adverse Childhood Experiences (ACE) Study. Am J Prev Med. 1998 May;14(4):245–58.

5. Bomysoad RN, Francis LA. Adverse Childhood Experiences and Mental Health Conditions Among Adolescents. J Adolesc Health Off Publ Soc Adolesc Med. 2020 Dec;67(6):868–70.

6. Barnett ML, Scheldrick RC, Liu SR, Kia-Keating M, Negriff S. Implications of adverse childhood experiences screening on behavioral health services: A scoping review and systems modeling analysis. Am Psychol [Internet]. 2021 Mar [cited 2024 Oct 14];76(2). Available from: https://pubmed.ncbi.nlm.nih.gov/33734801/

7. Loveday S, Hall T, Constable L, Paton K, Sanci L, Goldfeld S, et al. Screening for Adverse Childhood Experiences in Children: A Systematic Review. Pediatrics. 2022 Feb 1;149(2):e2021051884.

8. Correa MJ. Screening for adverse childhood experiences in the pediatric population in a clinical setting: a scoping review. 2024 Oct 16 [cited 2024 Oct 16]; Available from: https://osf.io/fs5pe/

9. Rethlefsen ML, Kirtley S, Waffenschmidt S, Ayala AP, Moher D, Page MJ, et al. PRISMA-S: an extension to the PRISMA Statement for Reporting Literature Searches in Systematic Reviews. Syst Rev. 2021 Jan 26;10(1):39.

10. PubMed PubReMiner: a tool for PubMed query building and literature mining [Internet]. [cited 2024 Oct 16]. Available from: https://hgserver2.amc.nl/cgi-bin/miner/miner2.cgi

11. Home - MeSH - NCBI [Internet]. [cited 2024 Oct 16]. Available from: https://www.ncbi.nlm.nih.gov/mesh/

12. Rayyan – Intelligent Systematic Review - Rayyan [Internet]. 2021 [cited 2024 Oct 16]. Available from: https://www.rayyan.ai/

13. Tricco AC, Lillie E, Zarin W, O’Brien KK, Colquhoun H, Levac D, et al. PRISMA Extension for Scoping Reviews (PRISMA-ScR): Checklist and Explanation. Ann Intern Med. 2018 Oct 2;169(7):467–73.

14. Gilgoff R, Koita K, Gentile B, Marques S. Adverse Childhood Experiences, Outcomes, and Interventions - PubMed [Internet]. 2020 [cited 2024 Oct 13]. Available from: https://pubmed.ncbi.nlm.nih.gov/32122559/

15. Bora N, Jones TR, Salada K, Brummel M. Inter-Clinician Variability in Primary Care Providers’ Adverse Childhood Experience Knowledge, Training, Screening Practices, and Perceived Intervention Barriers: an Exploratory Cross-Sectional Study. J Child Adolesc Trauma. 2021 May 20;15(2):285.

16. Kia-Keating M, Barnett ML, Liu SR, Sims GM, Ruth AB. Trauma-Responsive Care in a Pediatric Setting: Feasibility and Acceptability of Screening for Adverse Childhood Experiences. Am J Community Psychol. 2019 Dec;64(3–4):286–97.

17. Watson CR, Young-Wolff KC, Negriff S, Dumke K, DiGangi M. Implementation and Evaluation of Adverse Childhood Experiences Screening in Pediatrics and Obstetrics Settings. Perm J. 2024 Mar 15;28(1):180–7.

18. Negriff S, DiGangi M, Sidell M, Liu J, Coleman K. Assessment of Screening for Adverse Childhood Experiences and Receipt of Behavioral Health Services Among Children and Adolescents - PubMed [Internet]. 2024 [cited 2024 Oct 13]. Available from: https://pubmed.ncbi.nlm.nih.gov/36534401/

19. McBain RK, Levin JS, Matthews S, Qureshi N, Long D, Schickedanz AB, et al. The effect of adverse childhood experience training, screening, and response in primary care: a systematic review. EClinicalMedicine. 2023 Nov;65:102282.

20. Hillyer A. Implementation of Pediatric Adverse Childhood Experiences Screening in Primary Care. Dissertations [Internet]. 2021 Jul 8; Available from: https://irl.umsl.edu/dissertation/1072

21. Liu SR, Grimes KE, Creedon TB, Pathak PR, DiBona LA, Hagan GN. Pediatric ACES assessment within a collaborative practice model: Implications for health equity. Am J Orthopsychiatry. 2021;91(3):386–97.

22. Koita K, Long D, Hessler D, Benson M, Daley K, Bucci M, et al. Development and implementation of a pediatric adverse childhood experiences (ACEs) and other determinants of health questionnaire in the pediatric medical home: A pilot study. PloS One. 2018;13(12):e0208088.

23. Marie-Mitchell A, Lee J, Siplon C, Chan F, Riesen S, Vercio C. Implementation of the Whole Child Assessment to Screen for Adverse Childhood Experiences. Glob Pediatr Health. 2019;6:2333794X19862093.

24. Crichton KG, Cooper JN, Minneci PC, Groner JI, Thackeray JD, Deans KJ. A national survey on the use of screening tools to detect physical child abuse. Pediatr Surg Int. 2016 Aug;32(8):815–8.

25. Reading J, Nunez D, Torices T, Schickedanz A. A Qualitative Study of Pediatricians’ Adverse Childhood Experiences Screening Workflows. Acad Pediatr. 2022;22(8):1346–52.

26. Dube SR. Continuing conversations about adverse childhood experiences (ACEs) screening: A public health perspective. Child Abuse Negl [Internet]. 2018 Nov [cited 2024 Oct 15];85. Available from: https://pubmed.ncbi.nlm.nih.gov/29555095/

27. Bora N, Jones TR, Salada K, Brummel M. Inter-Clinician Variability in Primary Care Providers’ Adverse Childhood Experience Knowledge, Training, Screening Practices, and Perceived Intervention Barriers: an Exploratory Cross-Sectional Study. J Child Adolesc Trauma. 2022 Jun;15(2):285–96.

28. Sherfinski HT, Condit PE, Williams Al-Kharusy SS, Moreno MA. Adverse Childhood Experiences: Perceptions, Practices, and Possibilities. WMJ Off Publ State Med Soc Wis. 2021 Oct;120(3):209–17.

29. Jacob G, van den Heuvel M, Jama N, Moore Am, Ford-Jones L, Wong Pd. Adverse childhood experiences: Basics for the paediatrician. Paediatr Child Health [Internet]. 2019 Feb [cited 2024 Oct 15];24(1). Available from: https://pubmed.ncbi.nlm.nih.gov/30792598/

30. Campbell TL. Screening for Adverse Childhood Experiences (ACEs) in Primary Care: A Cautionary Note. JAMA. 2020 Jun 16;323(23):2379–80.

